# Safety and Efficacy of Dupilumab for the Treatment of Hospitalized Patients with Moderate to Severe COVID 19: A Phase IIa Trial

**DOI:** 10.1101/2022.03.30.22273194

**Authors:** Jennifer Sasson, Alexandra N. Donlan, Jennie Z. Ma, Heather M. Haughey, Rachael Coleman, Uma Nayak, Amy J. Mathers, Sylvain Laverdure, Robin Dewar, Patrick E. H. Jackson, Scott K. Heysell, Jeffrey M. Sturek, William A. Petri

**Author notes:** Corresponding author: William A. Petri Jr. University of Virginia, 345 Crispell Drive, Charlottesville Virginia 22908-1340, USA.

## Abstract

**Background:** A profound need remains to develop further therapeutics for treatment of those hospitalized with COVID-19. Based on data implicating the type 2 cytokine interleukin (IL)-13 as a significant factor leading to critical COVID-19, this trial was designed to assess dupilumab, a monoclonal antibody that blocks IL-13 and IL-4 signaling, for treatment of inpatients with COVID-19.

**Methods:** We conducted a phase IIa randomized double-blind placebo-controlled trial to assess the safety and efficacy of dupilumab plus standard of care versus placebo plus standard of care in mitigating respiratory failure and death in those hospitalized with COVID-19. Subjects were followed prospectively for 60 days. The primary endpoint was the proportion of patients alive and free of invasive mechanical ventilation at 28 days.

**Findings:** Forty eligible subjects were enrolled from June to November of 2021. There was no difference in adverse events nor in ventilator free survival at day 28 between study arms. However, for the secondary endpoint of mortality at day 60, subjects randomized to dupilumab had a higher survival rate compared to the placebo group (89.5% vs 76.2%, adjusted HR 0.05, 95% CI: 0.0-0.72, p=0.03). There were fewer subjects admitted to the ICU in the dupilumab group compared to placebo (33.3% vs 66.7%; adjusted HR 0.44, 95% CI: 0.09-2.09, p=0.30). Lastly, we saw downstream evidence of IL-4 and IL-13 signaling blockade in the dupilumab group through analysis of immune biomarkers over time.

**Interpretation:** Dupilumab was well tolerated and improved 60-day survival in patients hospitalized with moderate to severe COVID-19.

**Trial Registration:** This trial is registered with ClinicalTrials.gov, NCT04920916.

**Funding:** Virginia Biosciences Health Research Corporation, PBM C19, Henske Family Foundation, National Institutes of Health, National Cancer Institute

## INTRODUCTION

As in-hospital mortality remains at 10-26%^1,2^ paired with the ongoing threats of new SARS-CoV-2 variants, there remains a substantial need for additional therapeutics for those hospitalized with COVID-19. Current therapies against both the virus and with intention for immunomodulation have demonstrated variable and/or modest benefit. For example, the RECOVERY trial showed a mortality reduction from 26% to only 23% with dexamethasone use in those hospitalized with COVID-19 respiratory failure, with the greatest mortality benefit seen in those requiring mechanical ventilation at randomization^3^. Clinical trials for remdesivir, an antiviral nucleoside analog, have produced variable results, with the ACTT-1 trial demonstrating a 5 day reduction in clinical recovery time in those on supplemental oxygen^4^. Randomized controlled trials investigating interleukin (IL)-6 inhibitors have shown conflicting results, with some indicating a mortality benefit in those within 24 hours of intensive care unit (ICU) admission and others showing no difference in clinical outcomes between study groups^5,6^. Janus kinase inhibitors initially showed only a 1 day improvement in clinical recovery time when combined with remdesivir, with later trials since showing reduced mortality from 13% to 8% when combined with usual care in those requiring hospitalization and at least 1 elevated inflammatory marker^7,8^. Findings from these studies suggest a need for improvement in treatment of those admitted with COVID-19 pneumonia.

We have discovered that COVID-19 patients with high plasma IL-13 levels have a significantly greater risk of needing mechanical ventilation^9^. IL-13, which signals through the receptor IL-4Rα along with the closely related cytokine IL-4, is involved in eosinophilic inflammation, mucous secretion, goblet cell metaplasia and fibrosis, and has been regularly implicated in airway hyperresponsiveness and atopic disease^10^. We additionally found that neutralization of IL-13 in K18-hACE2 C57Bl/6J mice protected the animals from severe infection with SARS-CoV-2, as evidenced by reduced clinical score, weight loss and mortality^9^. The association of IL-13 along with other effectors of type 2 immunity with respiratory failure from COVID-19 has also been demonstrated in other observation studies^11,12^. These findings established mechanistic and biologic plausibility for IL-13 as a driver of pulmonary injury in COVID-19.

There are medications available to block IL-13 signaling: dupilumab, an anti-IL-4Rα monoclonal antibody, was approved for treatment of moderate to severe atopic dermatitis by the FDA in 2017. It reduces clinical severity in patients with allergic diseases including atopic dermatitis, asthma and chronic rhinosinusitis^13^. The original clinical trials demonstrated minimal adverse events with dupilumab use, favoring it as a steroid sparing therapy in atopic disease^14,15^. Post hoc analysis of initial studies saw reduced incidence of respiratory viral infections with its use^16^.

Dupilumab use was associated with greater survival from COVID-19 in retrospective analysis: using the TriNetX international electronic medical record (EMR) database, we previously identified a cohort of 350,004 patients with COVID-19, of whom 81 had been prescribed dupilumab prior to their COVID-19 diagnosis^9^. Patients on dupilumab had a 12.3% absolute risk reduction in mortality compared to a propensity score matched sub cohort of 81 patients with COVID-19 not on dupilumab but with atopic diseases for which dupilumab is routinely used^9^. Dupilumab has since been shown to reduce symptom severity and improve clinical outcomes in other observational studies utilizing large patient databases^17,18^.

The association of IL-13 with COVID-19 respiratory failure, the demonstration of survival benefit with IL-13 blockade in a mouse model and the retrospective EMR analysis showing reduced COVID-19 mortality in those receiving dupilumab for atopic disease, provided significant evidence for further exploration of dupilumab use for treatment of COVID-19. This along with the safety of dupilumab and the potential for a targeted approach to therapy led to the design of a clinical trial to test its use in those hospitalized with COVID-19.

## METHODS

### Design

This was a randomized, double-blind, placebo-controlled trial designed to assess the safety and efficacy of dupilumab use in 40 hospitalized patients from a single center with moderate to severe COVID-19 infection. It was approved by the University of Virginia Institutional Review Board (IRB) in June 2021 (NCT04920916). Eligible subjects were enrolled and randomized at a 1:1 ratio to receive either dupilumab or placebo, stratifying on disease severity measured by an oxygen requirement of ≤ 15 L/min or > 15 L/min by nasal cannula. Included were those over the age of 18 who were hospitalized with a positive reverse transcription polymerase chain reaction test (RT-PCR) for SARS-CoV-2 within the last 14 days and evidence of moderate to severe COVID-19 as defined by National Institutes of Health (NIH) COVID-19 Severity Categorization^19^. Patients requiring mechanical ventilation at the time of enrollment were excluded. Both arms received standard of care management per current NIH COVID-19 treatment guidelines, including dexamethasone and remdesivir as deemed appropriate by their primary provider^19^. Subjects received a loading dose of dupilumab (600 mg, given as two 300 mg subcutaneous injections) or placebo on day 0 with additional maintenance doses of 300 mg or placebo given on days 14 and 28 if the subject remained hospitalized and receiving active care^20^. Subjects were followed prospectively for 60 days.

### Outcomes

The primary outcome of the study was the proportion of patients alive and free of invasive mechanical ventilation at day 28. Safety outcomes were assessed via determination of the cumulative incidence of adverse events, including those previously reported to occur with dupilumab use (i.e., injection site reactions, eye/eyelid inflammation, conjunctivitis, herpes viral infection, eosinophilia)^20^. Additional clinical endpoints included all-cause mortality at day 28 and 60, proportion of patients alive and free of invasive mechanical ventilation at 60 days, hospital length of stay (LOS), ICU LOS, change in 8-point ordinal score and change in partial pressure of oxygen (PaO_2_) or oxygen saturation (SaO_2_) to fraction of inspired oxygen (FiO_2_) ratio. Plasma inflammatory markers, including C reactive protein (CRP), ferritin and a 47-plex cytokine panel were measured at various time points during the study. Additional type 2 inflammatory markers including TARC (CCL17), YKL40, eotaxin 3 (CCL26), arginase1 (Arg1), hyaluronan, soluble ST2 and total serum immunoglobulin E (IgE) were also measured. Ferritin, CRP and IgE levels were measured at the University of Virginia Clinical Laboratories while other biomarkers were measured by multiplex immunoassays or ELISAs depending on the analyte. SARS-CoV-2 baseline nucleocapsid (N)-protein level was measured from day 0, 2, 5, 7 and 14 available plasma of each subject using a microbead-based immunoassay, a highly sensitive detection method described in previous studies^21^. Day 0 nasopharyngeal (NP) swabs obtained for assessment of SARS-CoV-2 RNA positivity via RT-PCR underwent genomic sequencing to determine the SARS-CoV-2 lineage for samples with sufficient RNA using Artic v3 primers on either MiSeq (Illumina) or MinIon (Oxford Nanopore) using the and categorized according to PANGOLIN and World Health Organization^22,23^.

### Statistical Analysis

COVID-19 hospitalization data from UVA between March 2020 and April 2021 showed that 79.5% of COVID-19 inpatients were alive and free of mechanical ventilation at 28 days under usual care. With a pre-selected sample size of 40 patients and alpha=0.1 (one sided), we would be able to detect a difference of 17.7% in the proportion of subjects alive and free of mechanical ventilation at 28 days with 75% power.

Primary and secondary outcomes were analyzed under the intention-to-treat (ITT) principle. Safety outcomes were analyzed in the as treated population, including subjects who were enrolled and received at least one dose of study drug. Demographics, clinical and safety outcomes were analyzed initially with the Chi-square or Fisher’s exact tests for categorical measures and two-sample t-test or Wilcoxon rank sum for continuous measures, after assessment of normality. Treatment differences in ventilator free survival proportions were analyzed via logistic regression. Mortality differences were evaluated by the log-rank test and further in the Cox regression for time to death outcome. Baseline patient characteristics and known risk factors for severe disease in COVID-19, including age, sex, body mass index (BMI), comorbidities and COVID-19 vaccination status, were adjusted in regression models if initial analyses discovered imbalance in group characteristics^24^. Differences in the biomarkers between treatment groups were analyzed exploratively by t-test or Wilcoxon rank sum testing at each time point.

As an exploratory analysis, we included mechanical ventilation as a time varying variable in the Cox regression for further investigation of its influence on survivability. This allowed us to account for the significant change in mortality risk between pre- and post-intubation when a patient was placed on mechanical ventilation. We additionally tested differences in the likelihood of ICU admission between the two groups by the log-rank test. Lastly, after assessment of normality, N-protein levels were split into quartiles and analyzed by treatment group for influence on mortality via log-rank test and Cox regression. Regression models were adjusted for additional medications that were most likely to influence viral load, including monoclonal antibodies and remdesivir. Longitudinal N-protein levels over the first fourteen study days were evaluated by the treatment groups using the linear mixed effects models to account for within-subject correlations.

## RESULTS

### Patient and Virus Characteristics

Forty patients were enrolled from June 23, 2021 through November 11, 2021 (Fig S1). The groups were well matched with regard to age, BMI, race, ethnicity, comorbidities, vaccination status and days from COVID-19 symptom onset to enrollment (Table 1). Patients in the placebo arm were more likely to be male compared to the dupilumab arm (76.2% vs. 36.8%). There were no significant differences in non-study COVID-19 therapies received between the treatment groups (Table 1). Of those NP samples available for SARS-CoV-2 sequencing, 30 of 31 (96.8%) subjects had the delta variant and one subject in the placebo group had the iota variant (Table S1).

**Table 1:**
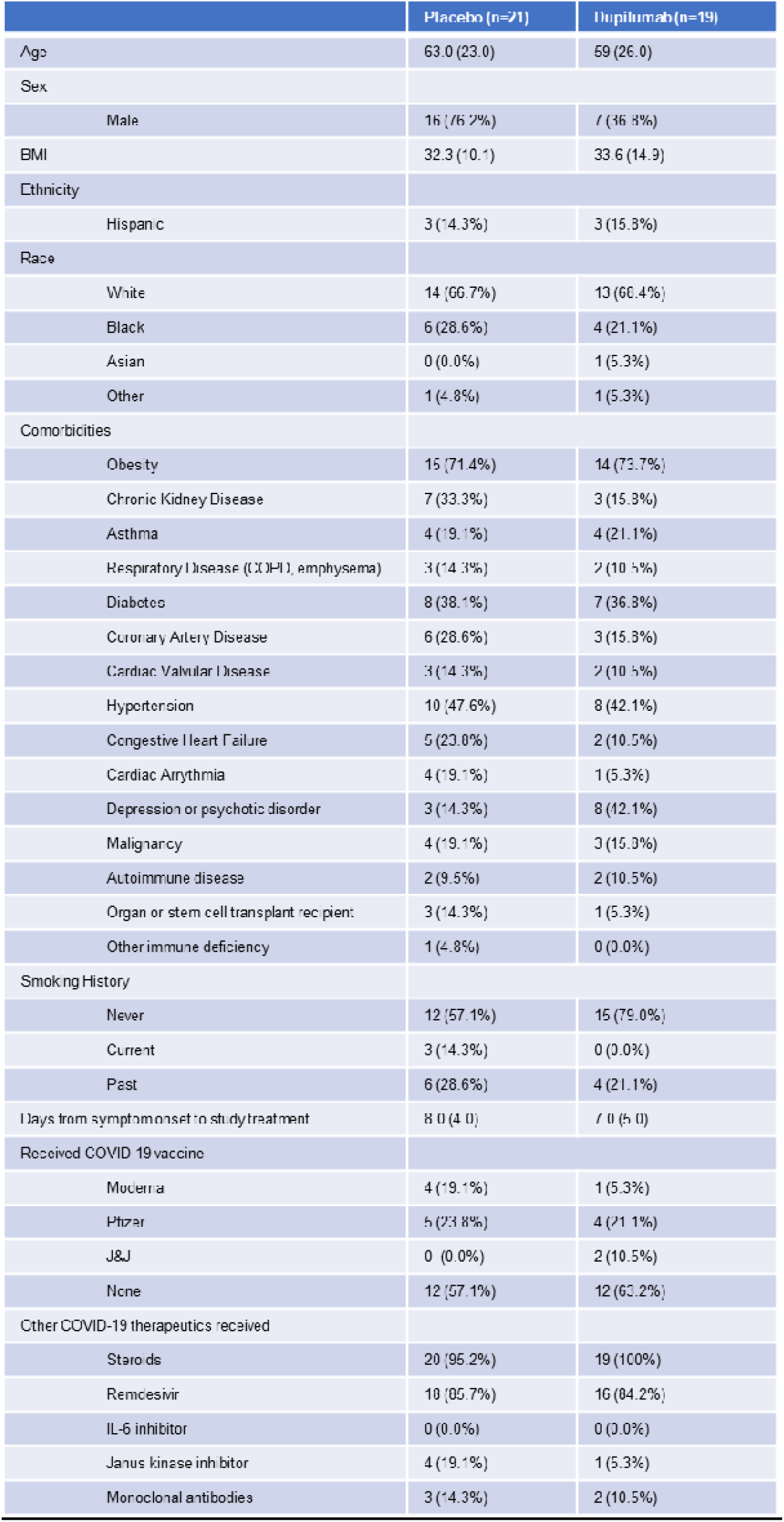
Patient characteristics. Continuous variables expressed as median (interquartile range). Categorical variables expressed as total n (percentage). Age expressed in years. Body Mass Index (BMI, kg/m^2^), Chronic Obstructive Pulmonary Disease (COPD), Johnson and Johnson (J&J).

### Safety

There were no significant differences in cumulative adverse events observed between the treatment groups (Table 2). In the dupilumab group, five subjects developed asymptomatic eosinophilia compared to one subject in the placebo group (Fisher’s exact p=0.09). There were no clinical consequences, including dermatologic, gastrointestinal, pulmonary, cardiac or neurologic, attributed to the peripheral eosinophilia seen in these subjects.

**Table 2:** Adverse events observed throughout the study period by treatment group. Other infections included *Clostridioides difficile* infection (1), bacteremia (2), urinary tract infections (2) and oral candidiasis (1). Categorical variables expressed as total n (percentage). Eosinophilia was defined as an absolute eosinophil count >0.6 k/uL at ≥ 1 measurement throughout the study period. *Difference between treatment groups was not statistically significant with Fischer’s exact p=0.09.

|  | Placebo (n=21) | Dupilumab (n=19) |
| --- | --- | --- |
| Injection site reactions | 0 (0.0%) | 0 (0.0%) |
| Conjunctivitis | 2 (9.5%) | 0 (0.0%) |
| Bacterial pneumonia | 1 (4.8%) | 2 (10.5%) |
| Herpes viral infection | 0 (0.0%) | 0 (0.0%) |
| Eosinophilia* | 1 (4.8%) | 5 (26.3%) |
| Hyper eosinophilic syndrome | 0 (0.0%) | 0 (0.0%) |
| Other infections | 2 (9.5%) | 4 (21.1%) |
| Cumulative | 6 | 11 |

### Clinical Efficacy

There was no significant difference in the primary endpoint of proportion of patients alive and free of mechanical ventilation at day 28 between the two groups (Table 3). However, by secondary endpoint at 60 days, 89.5% of subjects in the dupilumab group were alive compared to 76.2% for the placebo group as no patients remained on mechanical ventilation by day 60 in either group (Table 3). After adjustment for sex and mechanical ventilation as a time varying predictor, the risk of death over 60-day follow-up period was significantly lower in dupilumab group compared to placebo (Table 3; Fig 1).

**Table 3:** Primary and key secondary endpoints by treatment group. Primary endpoint was ventilator free survival by day 28. Secondary endpoints were ventilator free survival by day 60, mortality by day 60 and mortality by day 28. Proportions are listed as total n (%). The differences in the ventilator free survival proportions were evaluated using logistic regression, adjusted for sex. The differences in mortality risk were evaluated in the Cox regression, adjusted for sex and time varying mechanical ventilation.

|  | Placebo<br>(n=21) | Dupilumab<br>(n=19) | OR (95% CI) or HR (95% CI) | P value |
| --- | --- | --- | --- | --- |
| Proportion of patients alive and free of mechanical ventilation by day 28 | 18 (85.7%) | 15 (78.9%) | Unadjusted OR: 1.60 (0.31, 8.30) | 0.57 |
|  |  |  | Adjusted OR: 2.45 (0.40, 15.10) | 0.34 |
| Proportion of patients alive and free of mechanical ventilation by day 60 | 16 (76.2%) | 17 (89.5%) | Unadjusted OR: 0.38 (0.06, 2.22) | 0.28 |
|  |  |  | Adjusted OR: 0.44 (0.07, 2.96) | 0.40 |
| Mortality by day 28 | 3 (14.3%) | 1 (5.3%) | Unadjusted HR: 0.35 (0.04, 3.32) | 0.36 |
|  |  |  | Adjusted HR: 0.06 (0.003, 1.59) | 0.09 |
| Mortality by day 60 | 5 (23.8%) | 2 (10.5%) | Unadjusted HR: 0.40 (0.08, 2.05) | 0.27 |
|  |  |  | Adjusted HR: 0.05 (0.004, 0.72) | 0.03 |

**Figure 1:**
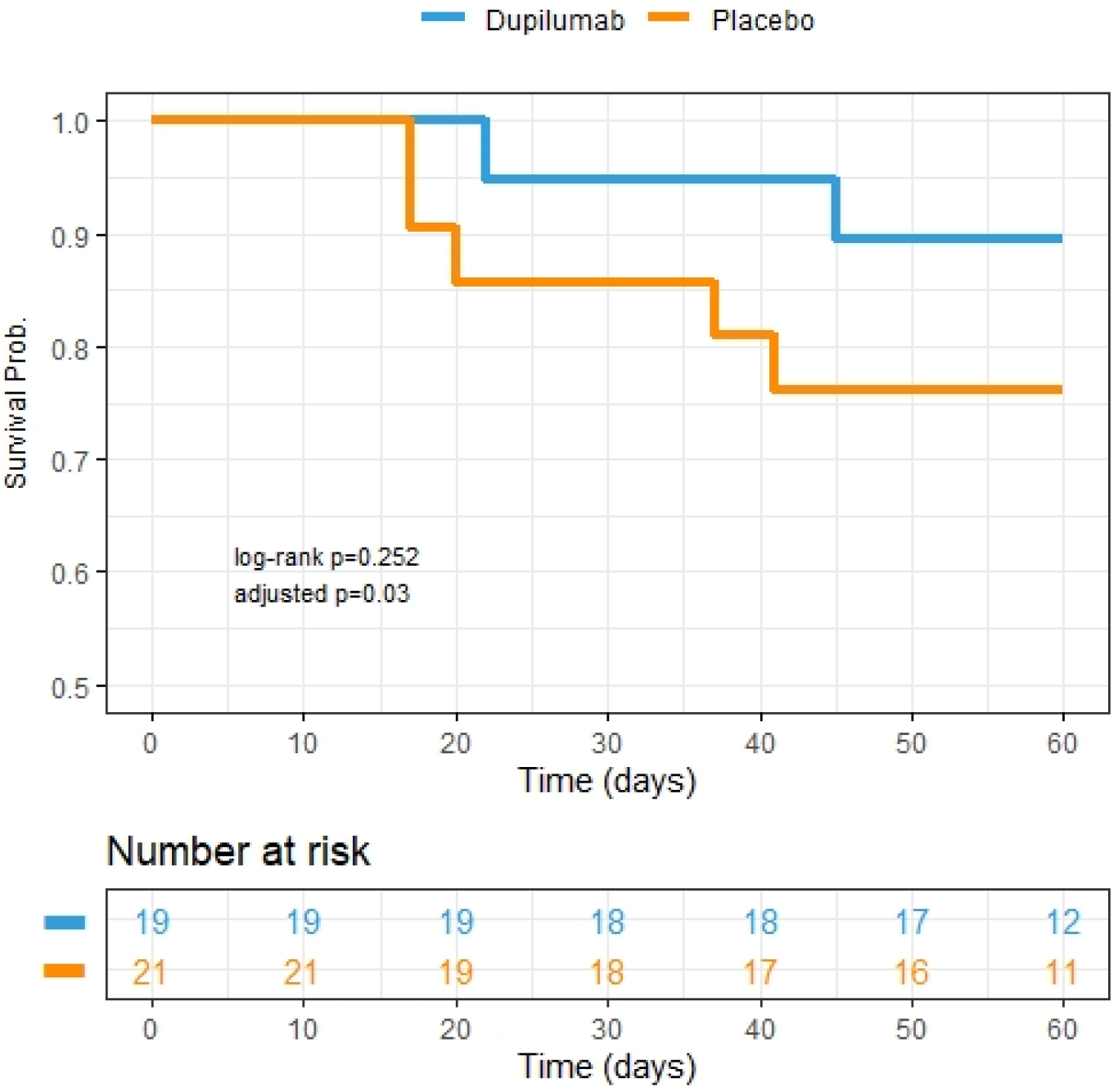
Kaplan Meier survival curves depicting 60-day mortality between two treatment groups. Dupilumab group is represented by blue line. Placebo group is represented by the orange line. Adjusted p value indicative of adjustment for sex and time varying ventilation in the Cox regression. Patient study visits occurred within an allotted range of exact study days and therefore the number at risk in the table is representative of patient data availability up until those exact days (i.e., if study visit was conducted on day 59 and no event had occurred, then the subject was included in the at-risk pool up until day 59 but not in that for day 60).

Numerically fewer subjects in the dupilumab group required ICU care (33.3%) compared to the placebo group (66.7%) though this difference was not statistically significant (log-rank p=0.23, HR 0.44, CI: 0.09-2.09, p=0.30 adjusted for sex, Fig 2). There was no difference in additional secondary endpoints between the two treatment groups (Table 4, Fig S2, Fig S3).

**Figure 2:**
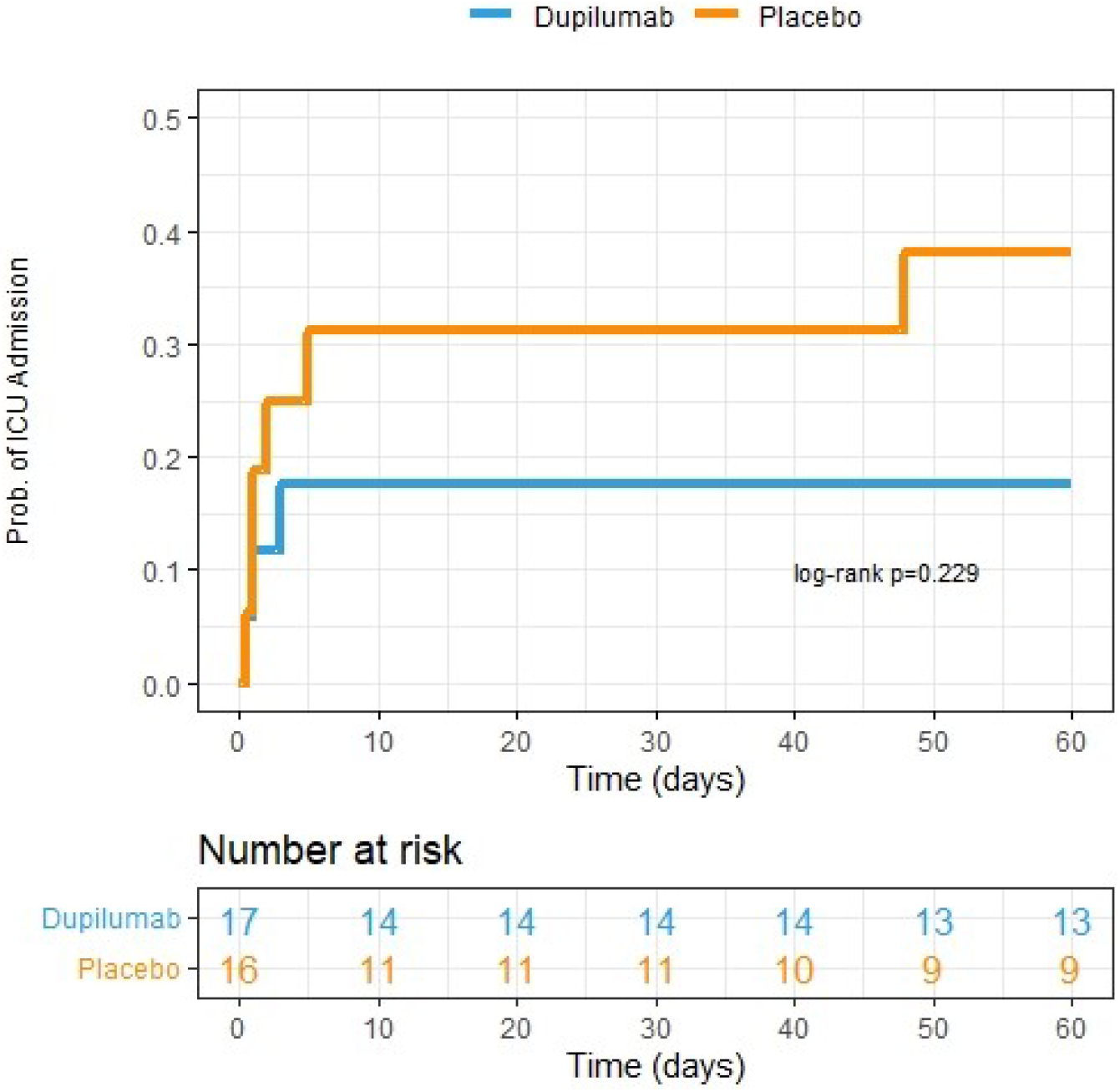
Kaplan Meier curve depicting need for escalation to ICU over 60-day study period. Patients already admitted to the ICU on day of enrollment (n=7) were excluded from analysis. Dupilumab group is represented by blue line. Placebo group is represented by the orange line. Patient study visits occurred within an allotted range of exact study days and therefore the number at risk in the table is representative of patient data availability up until those exact days (i.e., if study visit was conducted on day 59 and no event had occurred, then the subject was included in the at-risk pool up until day 59 but not in that for day 60).

### Biomarker Analysis

In both treatment groups, CRP, ferritin and IgE levels declined in the first two weeks with no significant difference in the change in measures from day 0 to 14 between groups (Fig S4). When looking at the change in absolute cell counts over time, there was an increase in eosinophils by day 14 in the dupilumab group compared to the placebo group (p=0.01 by Wilcoxon rank sum, Fig S5). Analysis of patient cytokine, chemokine and growth factors in serum at various study time points showed a decreased monocyte chemoattractant protein-1 (MCP-1) at day 7 in the dupilumab treatment group compared to placebo (p=0.04 by Wilcoxon rank sum, Fig S6). By day 14, there was a larger decrease in eotaxin-3 levels in the dupilumab group compared to the placebo (p=0.08 by Wilcoxon rank sum, Fig S6). Additionally, there was a trend towards decreased levels of YKL40 in the dupilumab group compared to the placebo by day 14 (p=0.26 by Wilcoxon rank sum, Fig S6). There was no statistically significant difference in baseline N-protein levels in the dupilumab group (median 671 ng/mL) compared to the placebo group (median 580 ng/mL; p=0.75 by Wilcoxon rank sum). When comparing the top quartile vs. the bottom three quartiles (i.e., bottom 75^th^ percentile) of baseline N-protein level within each treatment group, we found significant survival difference among the four groups (log-rank p=0.022, Fig S7). The 60-day mortality risk for those in the top quartile of baseline N-protein was 3.8 times of those in the bottom three quartiles after adjusting for treatment group, remdesivir use and monoclonal antibody use (95% CI: 0.78-18.7, p=0.098). N-protein levels in log-scale declined significantly from baseline to day 14 levels (p<0.0001), however, no difference was found in the rate of decline between the two treatment groups (p=0.17).

## DISCUSSION

In this randomized double-blind placebo-controlled trial, although there was no difference between study groups regarding the primary endpoint of 28-day ventilator free survival, the secondary endpoint of increased 60-day survival in the dupilumab group was achieved. Additionally, there were no safety signals seen with dupilumab use.

Although most deaths occurred in the placebo arm (5) compared to dupilumab (2), the overall mortality of subjects enrolled in this study (17.5%) was higher than expected, suggesting enrollment of a population with relatively higher disease severity. ICU mortality was 20% in the dupilumab group versus 36% in placebo, and ventilator mortality was 50% in the dupilumab group compared to 100% in placebo. Severity of illness seen in our study reflected that enrollment occurred during the delta surge and that the majority of those enrolled were unvaccinated, consistent with national data at the time^25^. For example, the National Hospital Care Survey (NHCS) data from the US Centers for Disease Control and Prevention (CDC), showed 11.9-13.1% in-hospital mortality in select hospitals throughout the United States during the month of August 2021 with ventilatory mortality rates ranging from 47.9%-74.1%, a time period during which this study enrolled subjects^26^. Furthermore, baseline N-protein levels were the same between the two groups and comparable to baseline N-protein levels of patients enrolled in the ACTIV3 trials^27^. As high N-protein levels are predictive of COVID-19 disease progression, a finding also demonstrated in this study, this suggests patients enrolled in our study were of comparable baseline disease severity^27^.

The detection of survival and mechanical ventilation differences at 60 days rather than at 28 days is consistent with reports of immunologic dysfunction from COVID-19 extending out to 8 months for mild to moderate COVID-19, with deaths from severe COVID-19 occurring out to 12 months^28,29^. Although the small size of our study limits broad conclusions about the mortality benefit of dupilumab, these findings combined support a late clinical benefit of blockade of a type 2 immune process in COVID-19. The response to dupilumab in asthma is also protracted with improvements in FEV1 first being observed 2 weeks after initiation of treatment^30^. Thus, the time to clinical effect of dupilumab in the acute COVID-19 setting may have limited our ability to see early clinical differences between the treatment groups. For example, subjects in our study who ultimately required mechanical ventilation did so within the first 8 days of the study, some within 1-2 days of enrollment, during a time in which drug concentration may have been lower, particularly in the context of a rapidly evolving clinical process.

Although biomarker trends seen in both groups were likely influenced by the steroids that almost all subjects received, we did see a reduction of the Type 2 immune markers YKL40 and eotaxin-3 in the dupilumab arm when compared to the placebo arm, indicative of the IL-4Rα blockade with inhibition of downstream mediators of the type 2 immune response. Increased peripheral eosinophil counts in the dupilumab group occurred by day 14, consistent with previous observations of dupilumab use in patients with atopic disease, likely due to decreased eosinophil uptake in tissue^30,31^. While we did not see IgE decrease at 2 weeks of dupilumab treatment, this is consistent with prior studies showing gradual decline of IgE levels compared to other biomarkers after dupilumab initiation^31^. We also saw reduction in MCP-1, a potent chemoattractant molecule of monocytes/macrophages, in the dupilumab group, high levels of which have been associated with COVID-19 disease severity^32,33^. Lastly, although recent invitro studies have shown that high IL-13 levels are associated with reduction in ACE2 receptor expression and decreased SARS-CoV-2 viral load, this is inconsistent with our study which shows similar rates of decline in N-protein levels in those who received IL-4Rα blockade compared to placebo^34^.

The study had several limitations. These included the lack of achievement of the primary endpoint of proportion of patients alive and free of mechanical ventilation at day 28, and the wide confidence intervals in the survival benefit of dupilumab at day 60. Additional limitations included unequal gender distribution between groups, patients were almost exclusively infected by the Delta variant of SARS CoV-2 and a higher-than-expected overall mortality rate.

The study also had several notable strengths including having as a foundation the preclinical data on the mechanism of disease exacerbation by IL-13 in COVID-19, originality in the study of type 2 immune inhibition, the use of a prospective placebo-controlled randomized and double-blind design, and demonstration of the safety of dupilumab. Importantly, there was evidence for mortality reduction and reduced ICU escalation with dupilumab use as we had predicted from animal models and retrospective human studies, despite sample size limitations. In light of the ongoing need for additional therapies for COVID-19 associated respiratory failure and the modest clinical benefits seen with other anti-viral and immunotherapies currently being used, the results of this study advance dupilumab as a promising treatment option for those hospitalized with COVID-19.

## Data Availability

All data produced in the present study are available upon reasonable request to the authors.

## ACKNOWLEDGEMENTS

We wish to thank the patients who consented to enroll in this study in an effort to help others with COVID-19 and the nursing staff and providers on the COVID-19 units for their assistance with the trial. We thank Jennifer White for IRB protocol preparation, Amy Warren and Lori Elder for IRB protocol preparation and IND preparation/submission and the staff at the investigational drug pharmacy at UVA. We’d also like to thank Mary Young and William Petri and the Biorepository and Tissue Research Facility at UVA who collected, organized and analyzed research samples. Lastly, we’d like to thank Helene Highbarger, Perrine Lallemand and Jeroen Highbarger of the Virus Isolation and Serology Laboratory, Frederick National Laboratory (FNL), and Ashley McCormack of Laboratory of Human Retrovirology and Immunoinformatics, FNL for their technical support on the nucleocapsid protein project.

## AUTHORS CONTRIBUTIONS

JS, AND and WAP contributed in funding acquisition; AND, JS, JZM and WAP contributed in conceptualization and methodology; HMH, RC, JS and WAP contributed in screening, enrollment and consent process; RC, HMH, UN, JS and WAP contributed to data collection and documentation; UN, RC, HMH and JS contributed in database management; HMH, RC, JS and WAP contributed in project administration and coordination; JZM, AND, JS and WAP contributed in data analysis and visualization; JS contributed in writing of original draft; all authors contributed in data interpretation, review and editing; WAP contributed in supervision; AJM contributed in organization and collection of viral sequencing data; SL and RD contributed in methodology, conceptualization, organization and collection of nucleocapsid protein data.

## ETHICS APPROVAL

Study approval was obtained through the Institutional Board Review at the University of Virginia and through the Food and Drug Administration (FDA) Investigational New Drug application.

