## Supplemental Tables/Figures for "Safety and Efficacy of Dupilumab for the Treatment of Hospitalized Patients with Moderate to Severe COVID 19: A Phase IIa Trial"

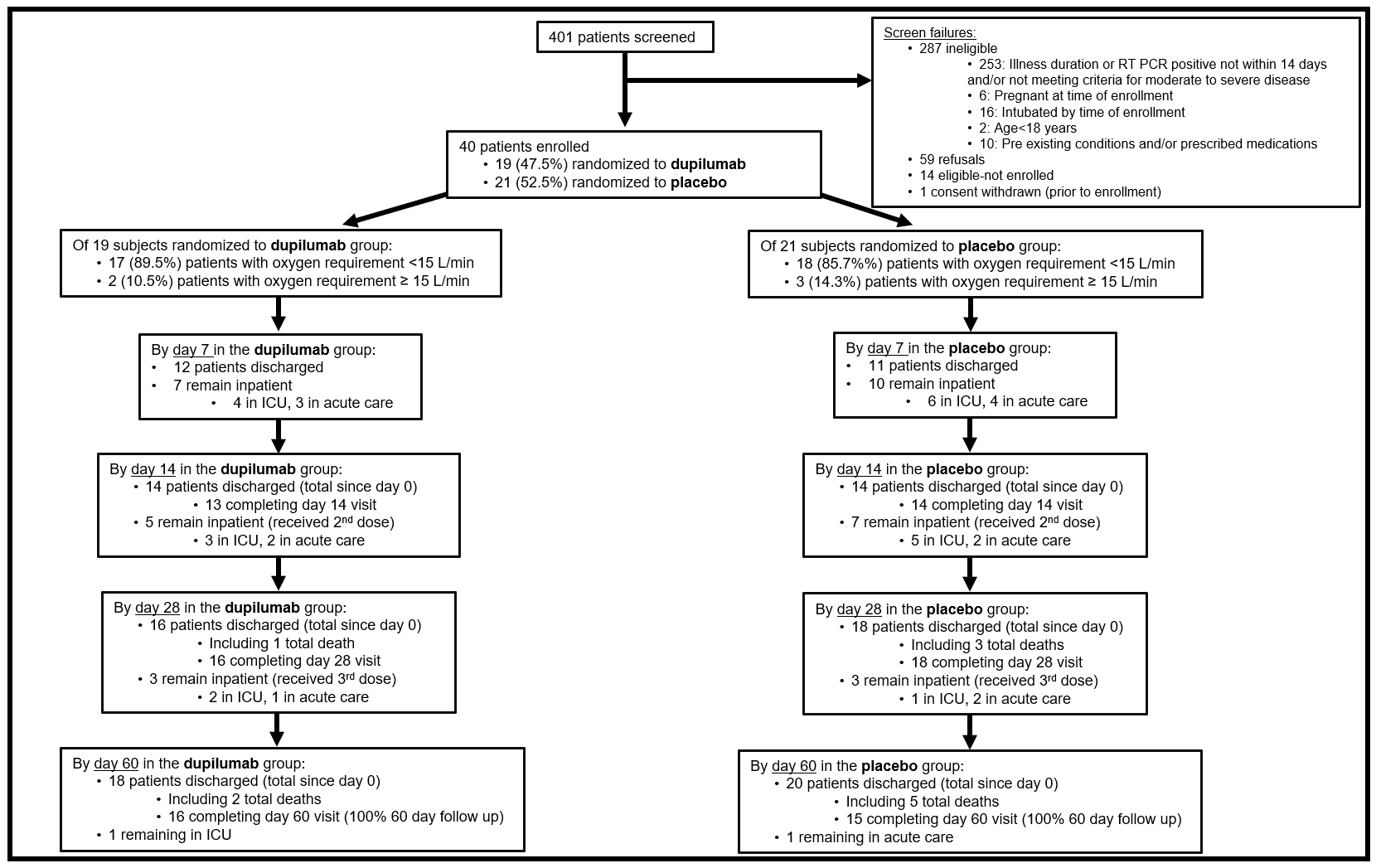

Fig S1: Study enrollment, randomization and timeline. Visit completion determined by clinical data collection and/or laboratory data collection by in person visits, telephone visits or data extraction from the electronic medical record.

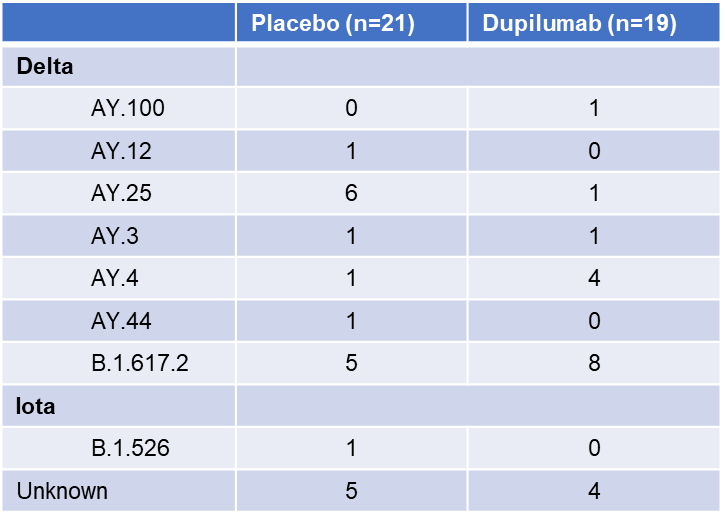

Table S1: World Health Organization (WHO) variant designations with Pango lineages as subheadings. Frequency of each lineage in study population listed by treatment group of those nasopharyngeal samples that were able to be sequenced.

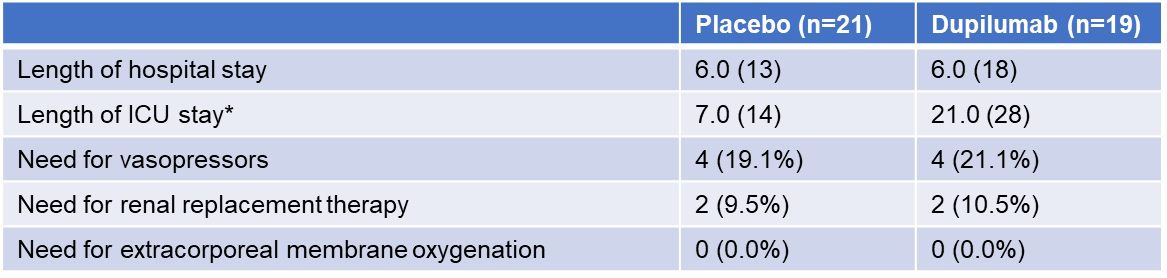

Table S2: Secondary clinical end points by treatment group. Continuous variables expressed as median (interquartile range). Categorical variables expressed as total n (percentage). Length of hospital and ICU stay measured in days. *Only includes patients requiring ICU admission during hospitalization (placebo n=11; dupilumab n=5).

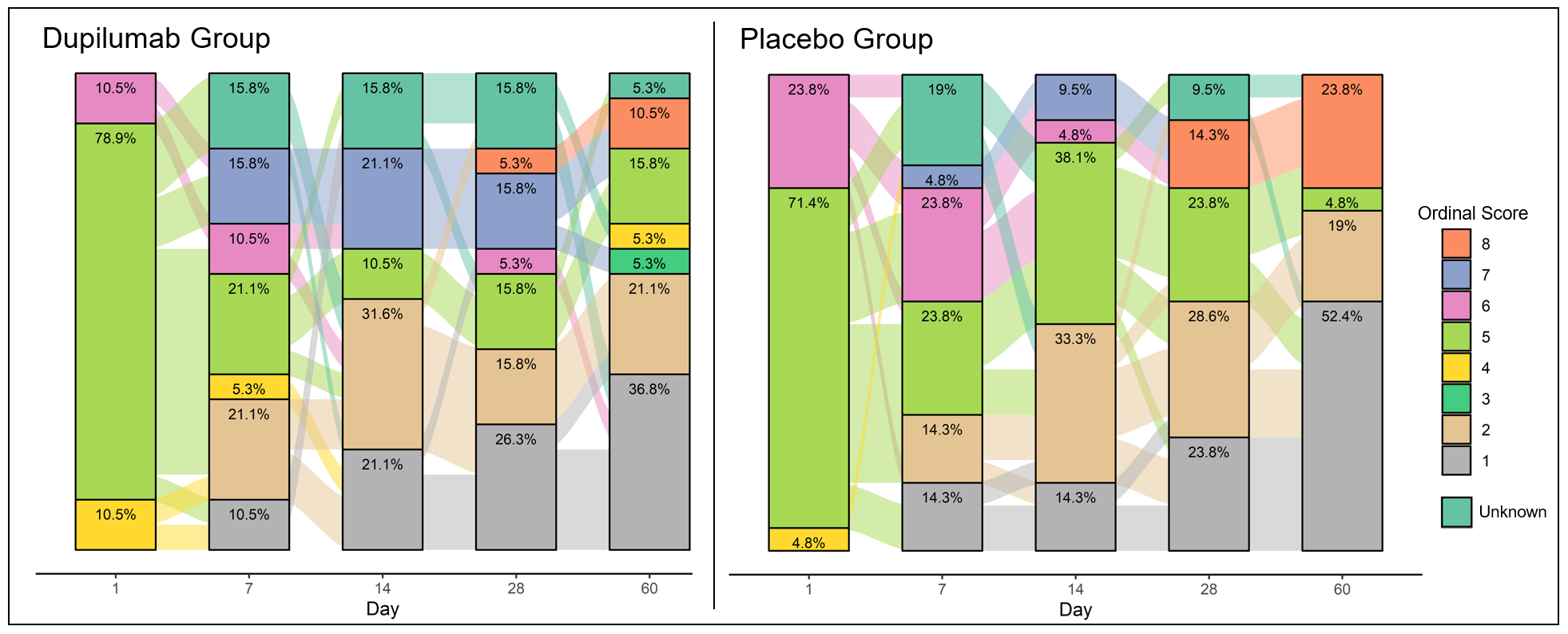

Fig S2: National Institute of Allergy and Infectious Diseases (NIAID) ordinal scores over time by treatment group. Proportion of patient ordinal scores at designated time points throughout the study period split into placebo (right) and dupilumab (left) groups. Group percentage for ordinal score listed within the colored box for each score. Ordinal scale as followed: 8) Death; 7) Hospitalized, on invasive mechanical ventilation or extracorporeal membrane oxygenation (ECMO); 6) Hospitalized, on non-invasive ventilation or high flow oxygen devices; 5) Hospitalized, requiring supplemental oxygen; 4) Hospitalized, not requiring supplemental oxygen - requiring ongoing medical care (COVID-19 related or otherwise); 3) Hospitalized, not requiring supplemental oxygen - no longer requires ongoing medical care; 2) Not hospitalized, limitation on activities and/or requiring home oxygen; 1) Not hospitalized, no limitations on activities.

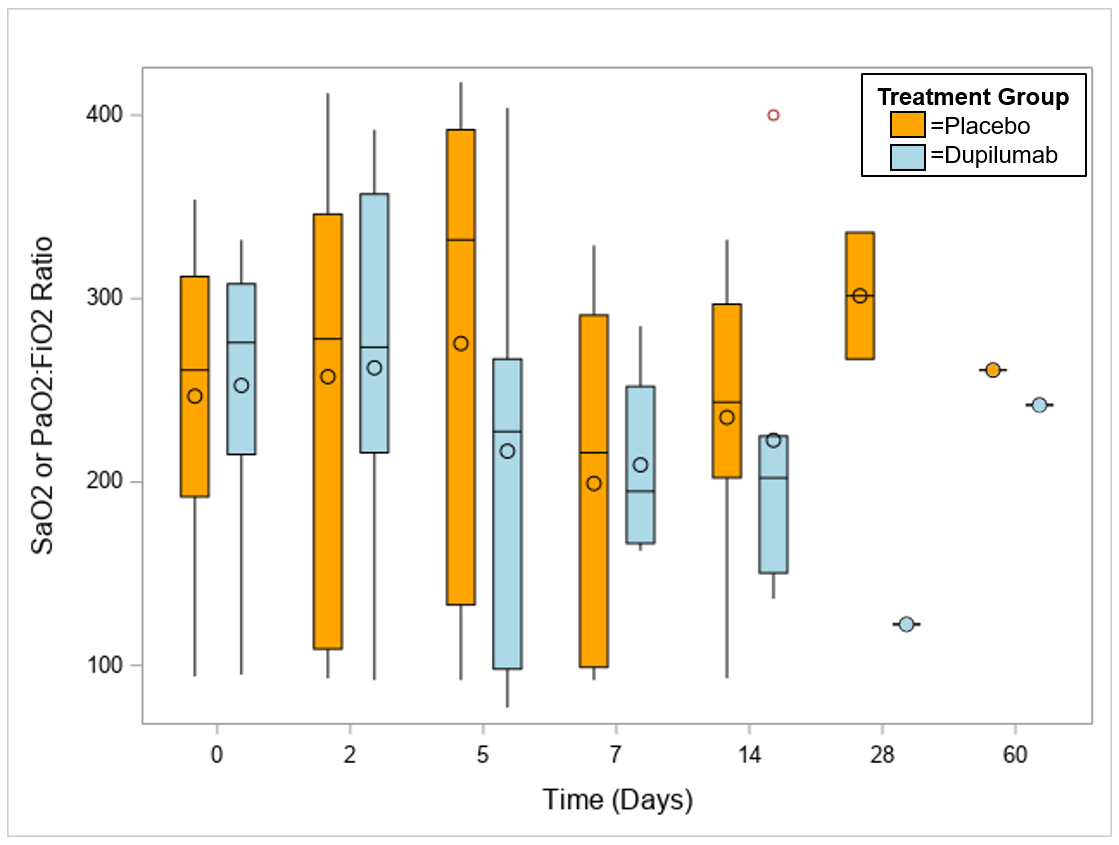

Fig S3: Box and whisker plots of SaO_2_:FiO_2_ Ratio or PaO_2_:FiO_2_ ratios over time by treatment group. Median ratios at each time point designated as horizontal line within boxes. Mean ratios depicted as open circles within boxes and filled circles outside of boxes. Dupilumab group represented by blue boxes/circles. Placebo group represented by orange boxes/circles.

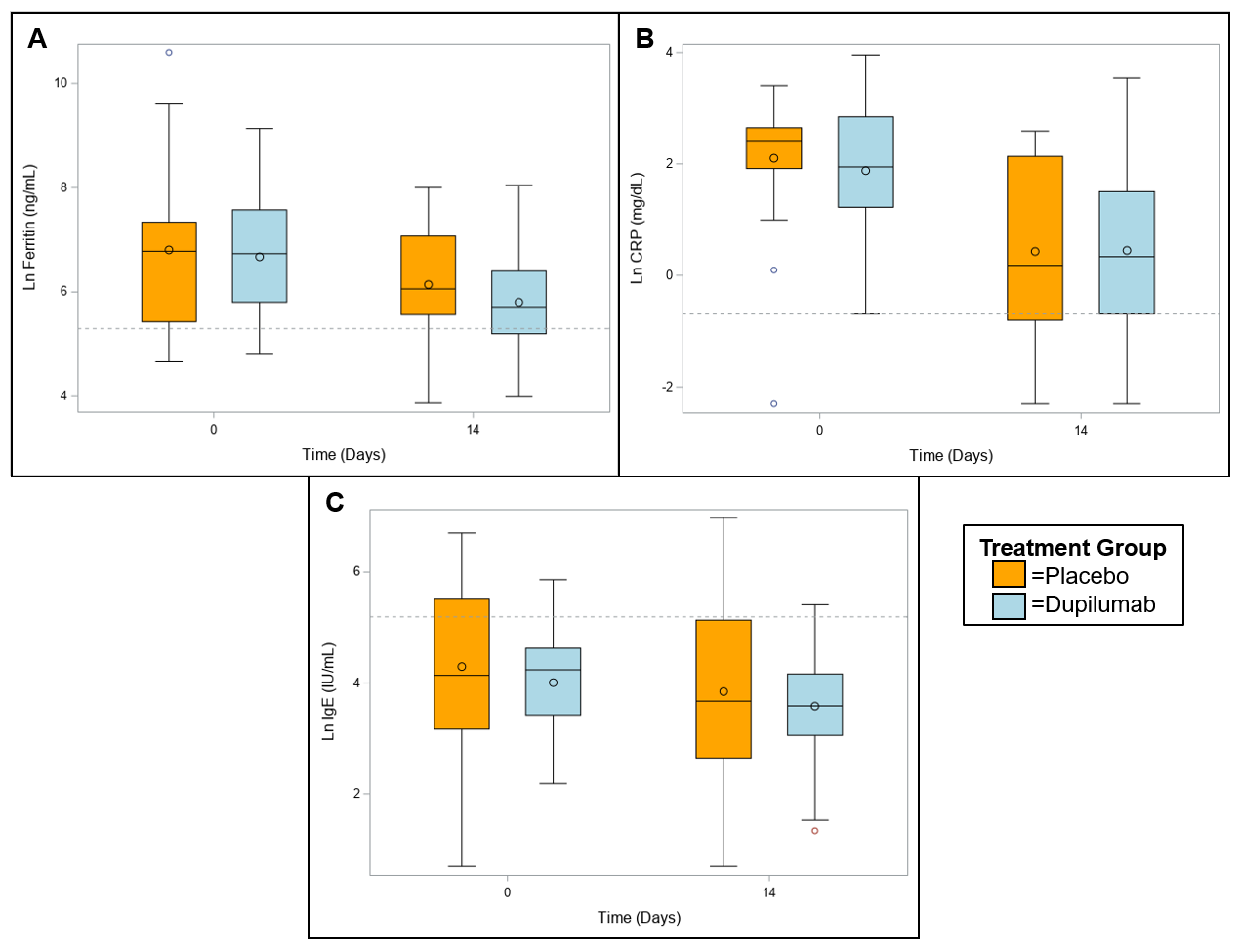

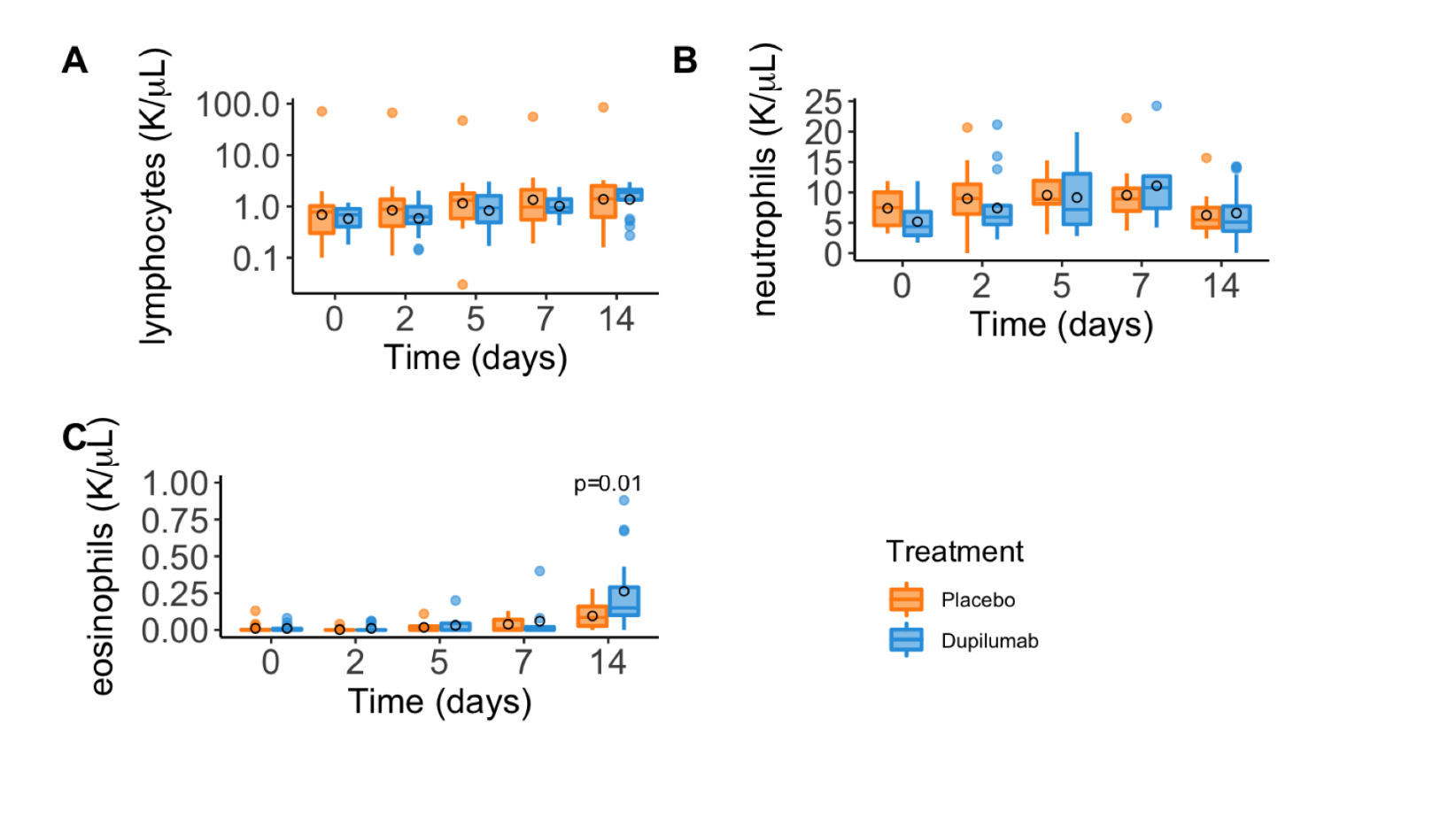

Fig S4: Box and whisker plots of ferritin (ng/mL, panel A), C-reactive protein (CRP, mg/dL, panel B) and immunoglobulin E (IgE, IU/mL, panel C) levels on study day 0 and study day 14 by treatment group. Values are natural log (Ln) transformed. Horizontal dotted line indicative of upper limit of normal (natural log transformed) for measurement via clinical lab. Solid horizontal line within box representative of median value and open circle within box representative of mean value. Blue boxes indicate dupilumab group and orange boxes indicated placebo group.

Fig S5: Box and whisker plots of absolute lymphocyte (K/UL, panel A), neutrophil (K/µL, panel B) and eosinophil (K/ µL, panel C) counts at study days 0, 2, 5, 7 and 14. Median value depicted by horizonal bar within box and mean value depicted by open black circle. Orange boxes indicate placebo group and blue boxes indicated dupilumab group.

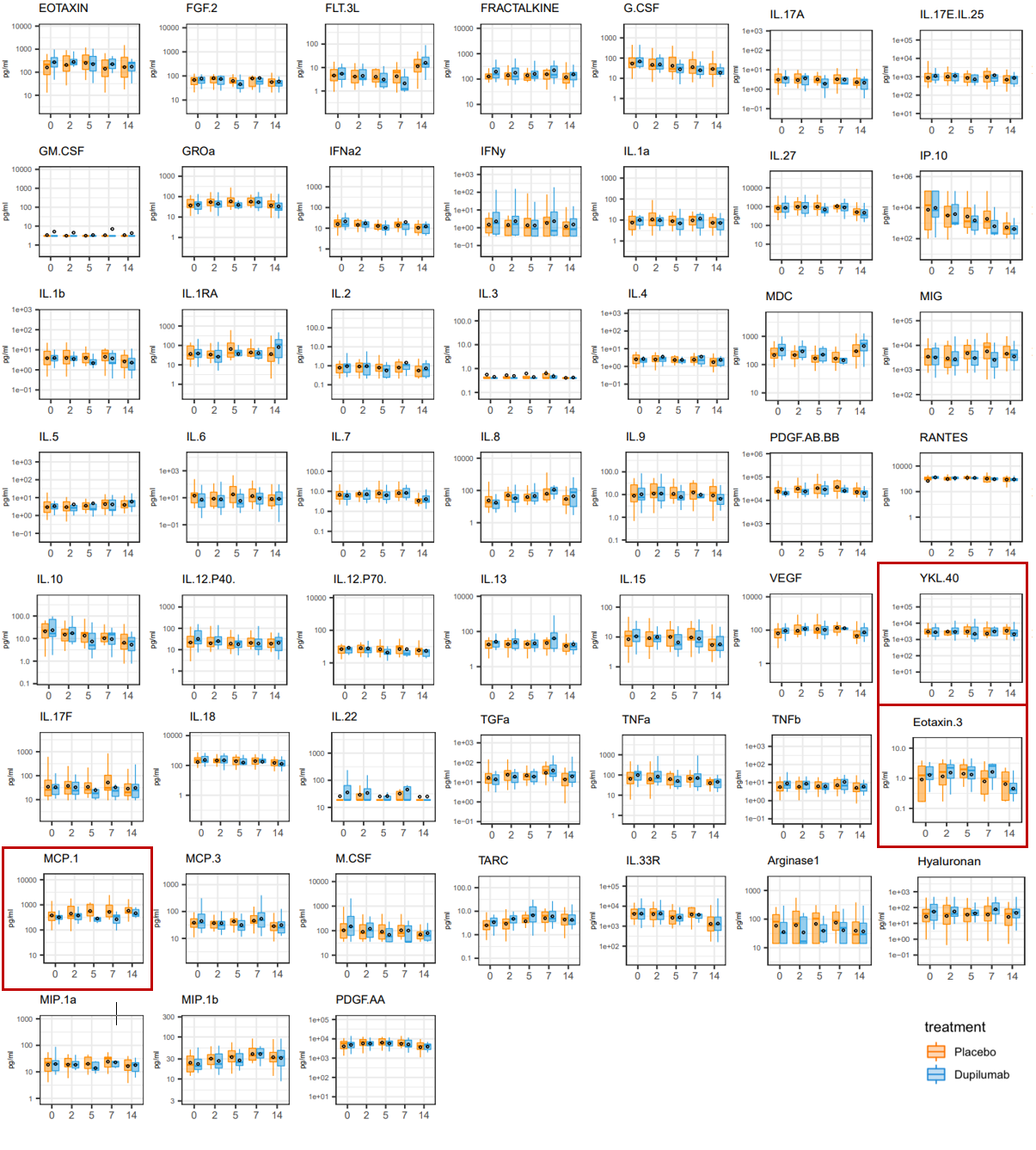

Fig S6: Box and whisker plots of cytokines, chemokines and growth factors in patient serum at study days 0, 2, 5, 7 and 14. Median value depicted by horizonal bar within box and mean value depicted by open black circle. Blue boxes represent the dupilumab group. Orange boxes represent the placebo group.

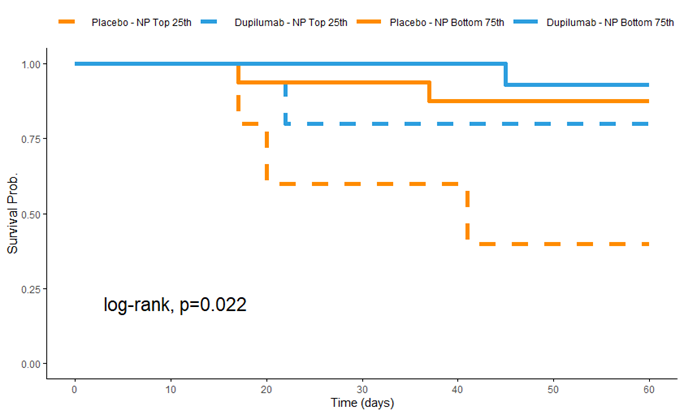

Fig S7: Kaplan Meier curve depicting 60-day survival probability by treatment group and N-protein (NP) quantile. Subjects in the dupilumab arm whose baseline N-protein level was in the top 25^th^ percentile of the group are represented by the blue solid line. Subjects in the placebo arm whose baseline N-protein level was in the top 25^th^ percentile of the group are represented by the orange solid line. Subjects in the dupilumab and placebo arms in the rest of the cohort (i.e., bottom 75^th^ percentile) are represented by the blue and orange dotted lines, respectively.
